# Repolarisation Speed May Vary with Characteristic Frequency in Human Spiral Ganglion Cells: Preliminary Observation from Electrically Evoked Compound Action Potentials

**DOI:** 10.64898/2026.04.23.26351590

**Authors:** Jason T.H. Lien, Stefan Strahl, Charlotte Garcia, Deborah Vickers

## Abstract

The human auditory system decomposes complex sounds into distinct components via a collection of processing steps. Knowing whether Spiral Ganglion Cells (SGCs) play an active role in the decoding of complex sounds can facilitate the development of Cochlear Implant (CI) coding strategies and clinical assessment tools. Early animal studies reported SGCs being similar across different characteristic frequencies (CFs). In this study, human electrically evoked compound action potentials (eCAPs) were analysed to probe the relationship between the reciprocal of CF and the duration of the eCAP. A significant relationship could indicate that SGCs may not simply be passive cables. eCAP datasets from 6 published studies (175 CI users, 1243 recordings) were analysed and their peaks were automatically labelled. The n1p2 latency was derived for each recording as a proxy of the action potential duration. The CF of each recording was estimated by mapping the average insertion angle of the electrode to the human SGC map. A weak but statistically significant relationship was observed between the n1p2 latency and the reciprocal of CF (random-effects model with random intercepts for subject, r = 0.09, p = 0.024, n= 450) supporting the hypothesis that lower CF is associated with slower repolarisation (longer n1p2 latency) in human spiral ganglion cells.

## Introduction

In the human auditory system, type I spiral ganglion cells (SGCs) connect the inner hair cells (IHCs) to the cochlear nucleus, marking the transformation from the mechanically transmitted sound into action potentials to encode information. The process of transmission begins at the periphery with sound sent to the cochlea from the outer and middle ears. The Basilar membrane that runs throughout the cochlea varies systematically in width and stiffness along its length resulting in a place of maximum displacement for different frequencies (Békésy & Wever, 1989; George Von Békésy, 1928). Outer hair cells (OHC) actively contract and expand corresponding to the cycle of the characteristic frequency (CF), which further amplifies and sharpens the response mechanically (Brownell et al., 1985; Rhode, 1978). IHC receptor potentials change periodically in line with the stimulus frequency, sending phase-locked signals to the auditory nerve (Johnson, 2015; Rose et al., 1967; Russell & Sellick, 1983), until they saturate. It is observed that the phase-locking activities have an upper-limit around 1500 to 10000 Hz (Verschooten et al., 2019a). These phase-locked signals provide additional information about frequency based on temporal firing. The auditory periphery codes frequency using both cochlear place and timing of neural firing.

With the rich information sent to the primary afferent system (ascending neural pathways), an important question emerges: Are SGCs simply passive cables, connecting the cochlea to the auditory nerve fibres, or do they play an active role in decoding the complex structure of sound?

Cochlear implants (CIs) are hearing devices provided to adults and children with severe-to-profound hearing loss. They bypass the auditory periphery and stimulate SGCs through electrical stimulation. The previous stages (e.g. filtering into bands) are included in the sound processing schemes which transform the signals into pulses for stimulating the SGCs. If SGCs are not just passive cables, the inherent characteristics could be utilised to provide salient temporal information. In addition, if SGCs have characteristics that are measurable, then CI stimulation approaches could be modified to take advantage of the natural SGC function along the cochlear length. This could be adapted for each individual based on their inherent neural function and measures of the viability of the electrode-neural interface (ENI; Lien et al., 2025). The ENI is the junction between the cochlea and the auditory nerve fibres which affects the current flow and how well the nerve fibres transmit information. It can be affected by insertion trauma which may cause SGC death and nerve atrophy and there could be issues due to nerve de-myelination. Electrode placement can affect current spread and interference between channels. This causes a great deal of variability in how well the auditory pathway processes the electrical stimulation (Bierer, 2010). Further understanding of SGC properties can help develop better ENI measures, which can lead to improvements in Cl performance.

Prior to data analysis, this study proposed the hypothesis that human SGCs vary systematically along the cochlea making the relationship between the reciprocal of the CF and repolarisation time, linear, with those tuned to higher CFs repolarising more rapidly than those tuned to lower CFs. If this relationship exists, each SGC can act as a low-pass filter ignoring (refractory period) stimulations with a rate higher than its corresponding CF.

The traditional view of SGCs being similar along the length of the cochlea emerged from pioneering studies reporting similar in-vivo post-stimulus time histograms despite different stimulating frequencies (Kiang et al., 1966; Sachs et al., 1974). With the advance of patch-clamping and in-vitro nurturing techniques, more recent rodent studies found that the distribution of potassium channels is related to the gangliotopical location, and SGCs with higher CF have a greater number of potassium channels (Adamson et al., 2002; Liu & Davis, 2007). This gradient of ionic channels is reflected by the recorded electrophysiological parameters such as action potential duration and stimulus to spike latency. Functionally, a gerbil study reported that high-spontaneous rate SGCs with lower CF rely on temporal coding to code the pure tone in noise (Huet et al., 2019). However, to the best of our knowledge, there hasn’t been a systematic exploration conducted with human SGCs. Electrically evoked compound action potentials (eCAPs) are neural telemetry approaches that utilise the implanted electrode array to stimulate a population of SGCs and record the compound response (Abbas et al., 1999; Brown et al., 1990; Charlet De Sauvage et al., 1983; Miller et al., 2000). A clear eCAP response is typically comprised of a valley (N1) and a peak (P2) happening before the first 1.5 milliseconds after the stimulus presentation.

In this study, human eCAP data sets collected from CI listeners were analysed to investigate the relationship between the estimated CF and recorded N1-P2 latency (the time needed for the potential to recover from valley to peak, reflecting repolarisation).

## Method

### Data Source

Raw eCAP recordings from 6 published studies were shared with us by the authors (Brill et al., 2009; Garcia et al., 2021, 2023, 2024; Garcia & Carlyon, 2025; van de Heyning et al., 2016). Among the studies, ethical approvals of the experiments were obtained from the institutional review board, research ethic committee, international Freiburger ethic commission, and/or other relevant ethical review committees (more details provided in-text of the above-cited articles). For each recording, the implant model, electrode number and time-amplitude data points were provided. Before data processing, the dataset comprised 131 MED-EL electrode users (Standard, n = 120; Flex, n = 11), 21 Cochlear electrode users (CI24RE, n = 4; CI512, n = 3; CI612, n = 1; CI422, n = 1; CI522, n = 5; CI622, n = 7), and 23 Advanced Bionics electrode users (Mid-Scala, n = 9; SlimJ, n = 8; 1J, n = 3; Helix, n = 3). In total, 1243 eCAP recordings were shared.

## Data Processing

### Automatic eCAP Labelling

To ensure unbiased judgement of the valley (n1) and peak (p2) of the waveform, an automatic labelling algorithm was developed. For each recording, the time-amplitude data points were used to perform ordinary least squares (OLS) fit of a custom function defined as follows:

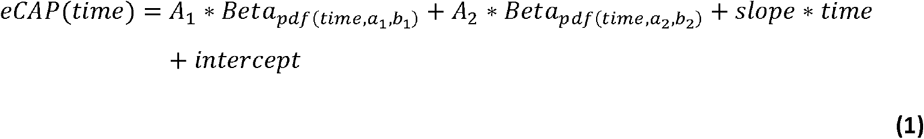

where *Beto*_*pdf*_ is the probability density function of the Beta distribution, *A*_*1*_ controls the size of the valley (first Beta PDF), *a*_*1*_ and *b*_*1*_ are the parameters of the first Beta PDF, controlling the location of the valley and the shape of it, *A*_*2*_ controls the size of the peak (second Beta PDF), *a*_2_ and *b*_*2*_ are the parameters of the first Beta PDF, controlling the location of the peak and the shape of it, *slope* and *intercept* provide space to model the noise caused by the residual current.

Prior to fitting, both the time and amplitude axes were scaled by dividing each by its respective maximum value to promote efficient convergence of the fitting algorithm. The “clean” eCAP was then obtained by removing the linear component (specifically, the slope and intercept terms) from the fitted function. Within this cleaned waveform, N1 was defined as the time point at which the eCAP reached its minimum value, and P2 as the time point at which it reached its maximum value. Figure 1 demonstrates an example of the labelling produced using this algorithm. The quality of the marking was quantified using the signal-to noise ratio (SNR) metric. Signal strength was defined as the difference between the P2 and N1 fit voltage values (A2 and A1, respectively, eCAP amplitude), and noise was defined as the mean of the largest 20 % of absolute errors between the recorded data points and the fitted function. For each automatic fit, the distribution of error sizes was mostly geometrically distributed. Therefore, averaged error does not reflect the fitness around the n1p2 region as well as the mean largest 20% absolute errors. The 20% cutoff was selected to differentiate fits that both have small errors around the eCAP tail but differ in fitness around the n1p2 region, with the 20% value selected based on visual inspection of the distribition of the errors and the eCAP waveforms. SNR was then defined as the ratio between the eCAP amplitude and noise.

**Fig. 1.**
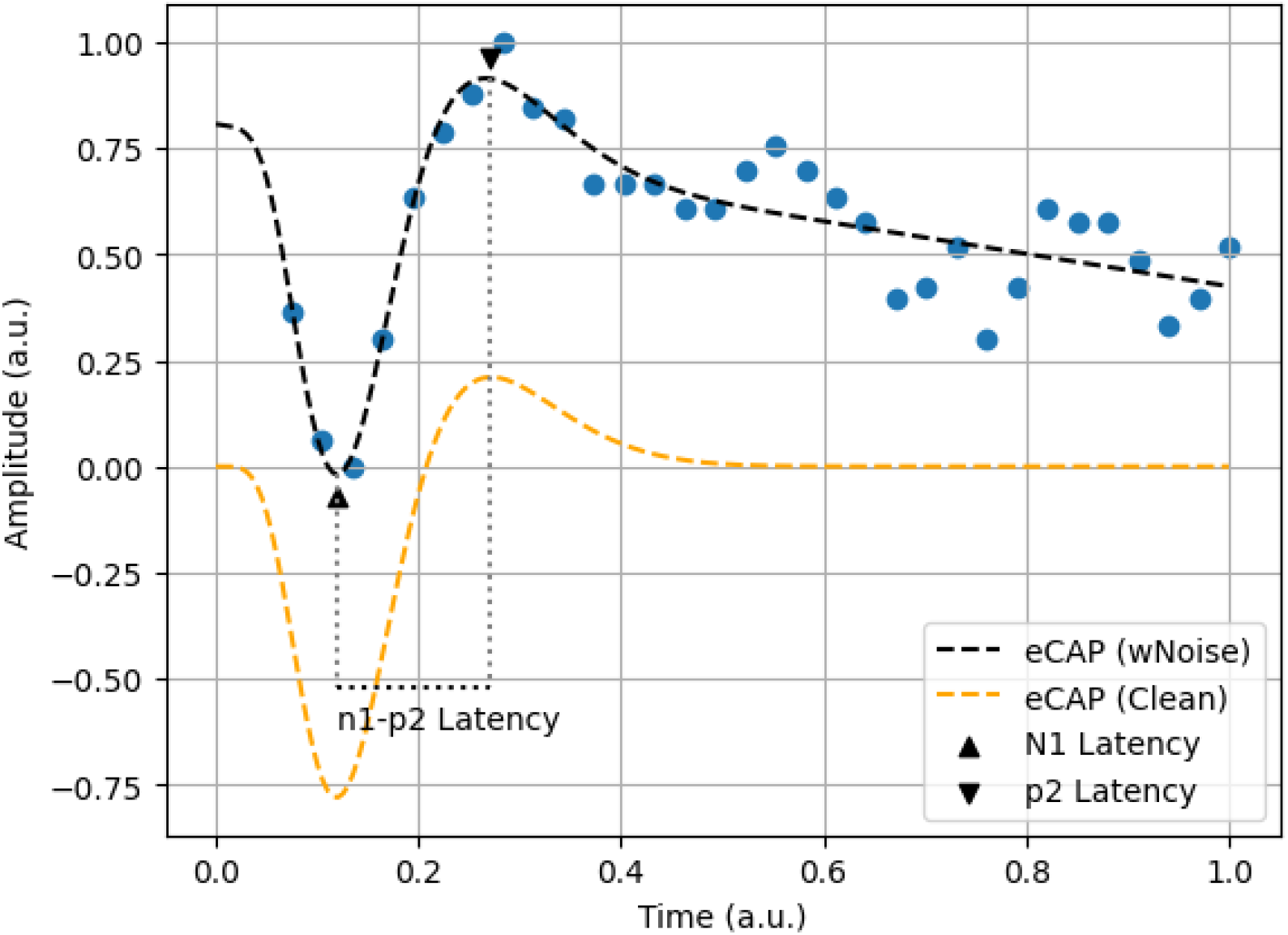
An Example of the Automatic eCAP Labelling. Blue dots mark the recorded data points. The black dashed line represents the line of best fit (with linear component). The orange dashed line represents the clean eCAP. The up and down triangles indicate the timings of N1 and P2 respectively. This example was from a Cochlear device (participant C19L, probe electrode number 4, recorded at electrode 6; Garcia et al., 2021).

### Characteristic Frequency Estimation

Insertion angle for each electrode array model has been reported by several published articles (Kırbaç et al., 2025; Landsberger et al., 2015; Peters et al., 2019; Sipari et al., 2024; Söderqvist et al., 2022; Wright et al., 2005). If a one-to-one, electrode-to-angle, relationship was given, this mapping was adopted directly. If not, the electrode-to-angle map was estimated from the electrode contact spacing information and the reported insertion angles of the most apical and basal electrodes from the articles. Given the model and electrode number of each recording, an insertion angle was assigned based on the results. CF is then estimated by mapping the insertion angle using the human SGC map (Stakhovskaya et al., 2007).

### Data Cleaning

Not every recording was included in the analysis stage. Different settings (stimulation level & artefact cancellation technique) were applied across recordings. As a result, if any given participant-electrode combination was measured more than one time, the setting that yielded the highest SNR was selected. To avoid bias introduced by the algorithm and poor recordings, only recordings with an SNR larger or equal to 4 were included. This SNR inclusion standard was determined by testing candidate standards at 0.5 SNR steps and the first author, a registered clinical audiologist, visually inspected the labelled eCAP recordings. This ratio was selected to ensure the automatic labels meet the typical judgement of the first author (registered clinical audiologist) consistently. The analysis was limited to data points with an estimated CF of 4000 Hz or lower. This is because the upper limit of phase-locking is thought to be around 1500 Hz to 10000 Hz (here we use 4000 Hz as the cutoff) (Verschooten et al., 2018, 2019a). After cleaning, 450 of the 1243 eCAP recordings, from 137 implants, were included in the analysis. Most exclusions were due to the frequency criterion, and approximately 10% of the remaining recordings were excluded based on the SNR criterion.

## Statistical Analysis

Fixed-effects and mixed-effects linear models, along with power calculations, were used to examine the relationship, using custom scripts written in Python3 with SciPy version 1.15.3, NumPy version 2.2.6, and Statsmodel version 0.14.4 (Harris et al., 2020; Seabold & Perktold, 2010; Virtanen et al., 2020). With the current automatic labelling algorithm, the smallest meaningful latency difference is approximately 293 μs (6 sampling points). If a meaningful latency difference between CF = 100 Hz and CF = 4000 Hz exists, the corresponding effect size for the 1/CF is approximately 30 (292.8/9.75 ≈ 30). Power analysis indicated that, under the fixed-effects model, an effect of 1/CF of approximately 37 would be detectable with 80% power at α = 0.05. All scripts used for this study are available in the supplementary code section.

## Results

Figure 2 shows the trend between n1p2 latency and CF. The median n1p2 latency of each location followed the hypothesised linear relationship between the reciprocal of CF and n1p2 latency. To control for differences across studies and subjects, a random-intercept (for subject) mixed-effects linear model indicated that 1/CF significantly predicted n1p2 latency (β = 22.08, SE = 9.77, r = 0.09, p = 0.024; marginal R^2^ = 0.008, conditional R^2^ = 0.49, 137 subjects, 450 observations). The marginal R^2^ reports the proportion of variance explained by 1/CF alone (r-value is then derived from the positive square root of it) and the conditional R^2^ indicates the goodness-of-fit of the mixed-effects model (Nakagawa & Schielzeth, 2013). Figure 3 shows individual data points and the line of best fit. Prior to the mixed-effects model, a simple fixed-effect model was used to investigate the relationship. However, the residuals showed patterns of autocorrelation. Thus, the result of an OLS regression with cluster-robust standard error estimation is provided in the following for completeness but we don’t consider the result fully representative of the nature. The regression model indicated that 1/CF was positively associated with n1p2 latency (β = 49.44, SE = 14.48, t = 3.42, p = 0.001). The model explains a modest proportion of variance (R^2^ = 0.04, r = 0.2; N = 450).

**Fig. 2.**
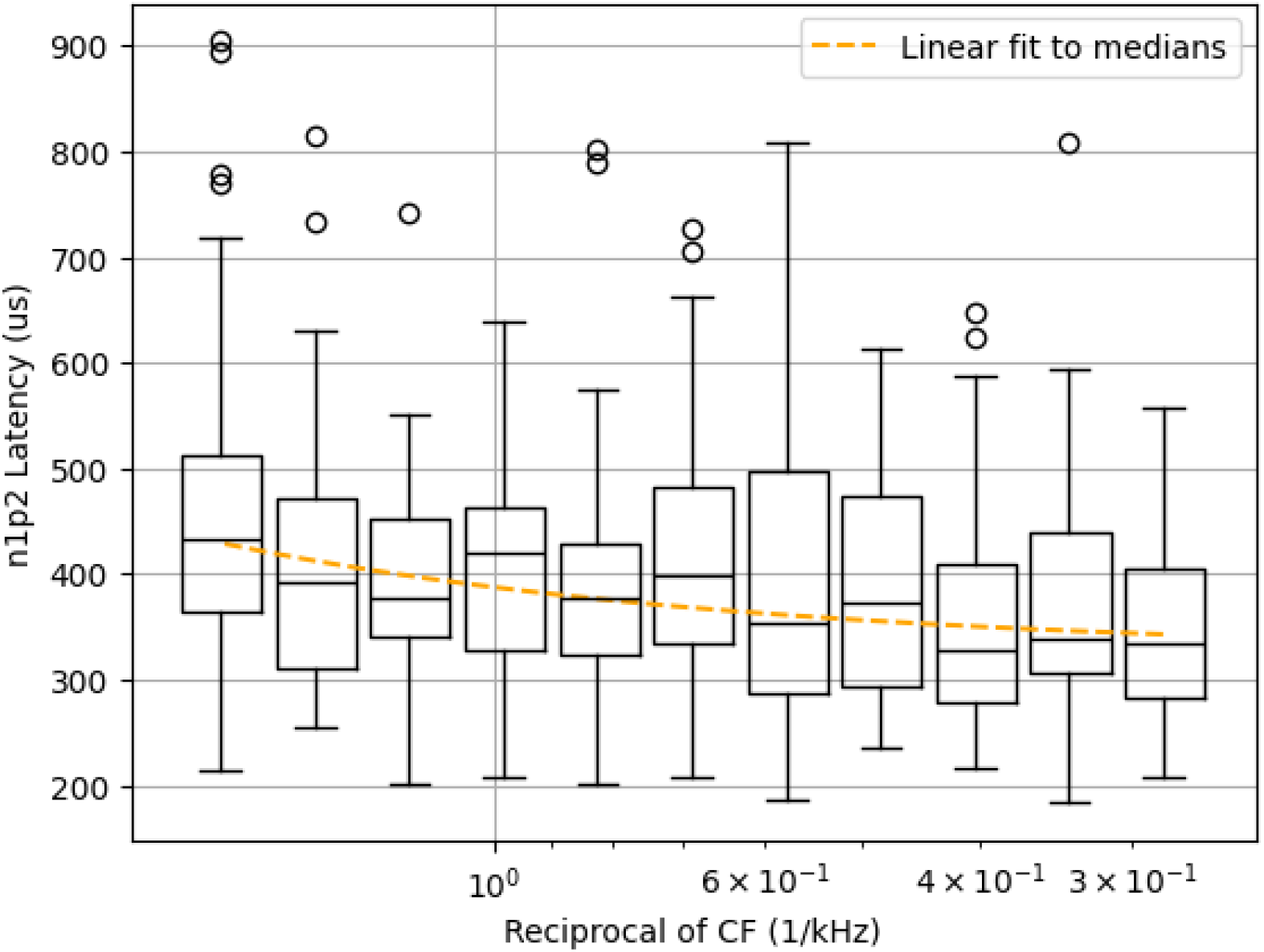
General Trend. Boxplots summarise n1p2 latencies across CF bins (x-axis), with the central line indicating the median, boxes representing the interquartile range, and whiskers extending to 1.5 times the interquartile range. Open circles denote outliers.

**Fig. 3.**
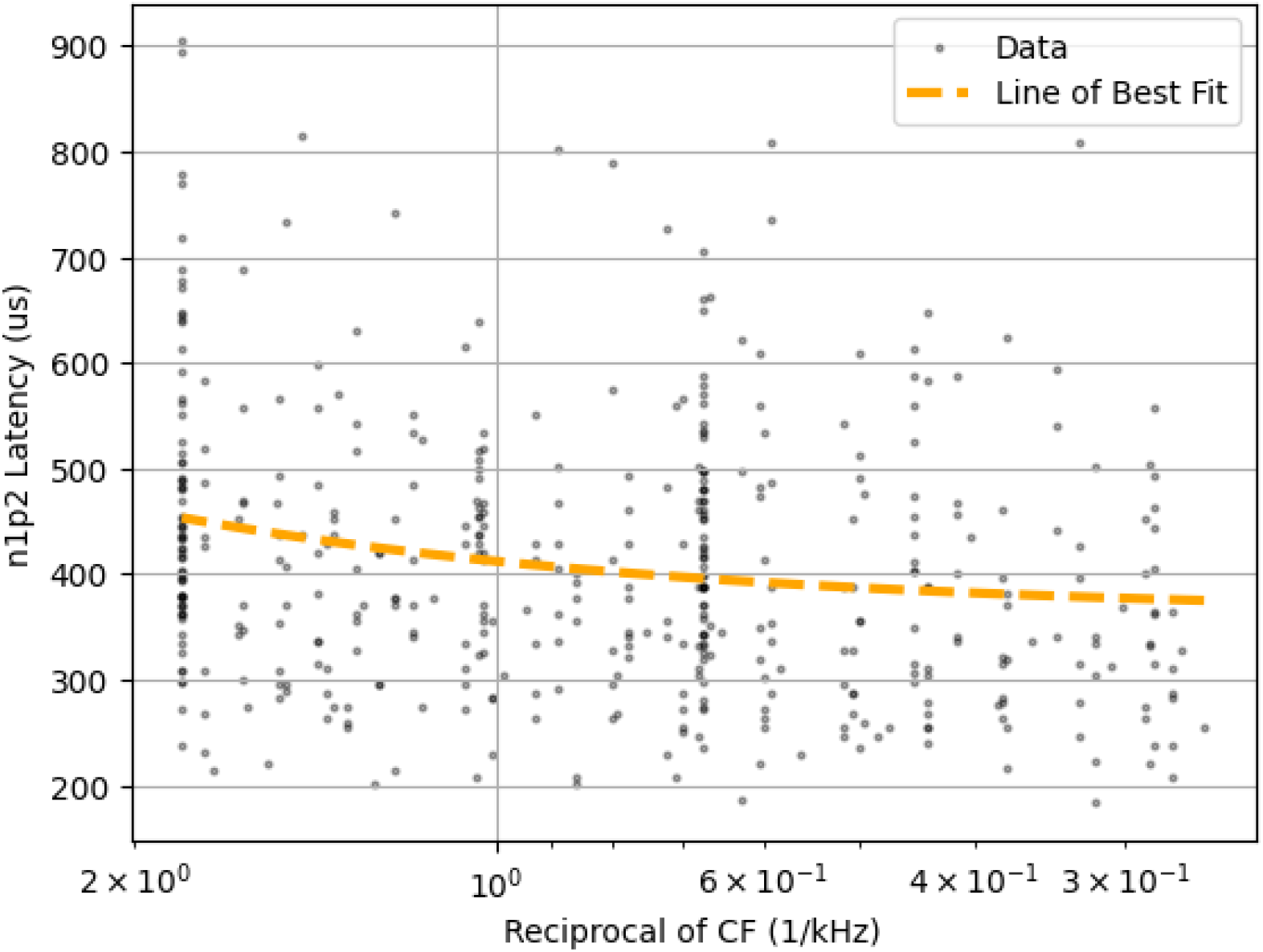
OLS Regression. Each semi-transparent black dot represents one data point. The orange dashed line represents the line of best fit.

## Discussion

### Spiral Ganglion Cells Differ along the Cochlea

To the best of our knowledge, this study is the first to report the relationship between CF and n1p2 latency, a proxy of action potential duration, using human in-vivo data. Previous animal studies reported that there is a systematic variation in the number of potassium channels related to the cochlea location (Adamson et al., 2002; Liu & Davis, 2007). Such gradient of ionic channels lay the foundation for different repolarisation speeds, which was reflected by the recorded action potential duration and stimulus to spike latency. These findings, together from in-vitro animal studies and the current in-vivo human study, support the hypothesis that SGCs are not simply passive cables. SGCs with higher CF, functionally, need to repolarise faster to encode temporal information via the spike timing. This characteristic of SGCs is similar to the previous stages of the peripheral auditory nerve system, building a hierarchy of filters to refine the complex input signal into separated components. The coherent findings across species might indicate that SGCs, in general, differ purposely along the cochlea.

### Limitations

The finding of this study was limited by several factors, estimated CF based on published averages for different electrode array types, unmeasured individual auditory nerve status and different eCAP recording settings (number of sweeps & artefact cancellation method). The current method of mapping device to insertion angle did not take into account individual factors that could affect the results, and this inherently causes a great deal of variability in responses. The insertion depth can vary greatly between different CI users, even for the same model of device and hence the stimulated SGC populations could be different CFs from the ones that we estimated based on average insertion depths reported in the literature (Canfarotta et al., 2020; Kirbaç et al., 2025; Landsberger et al., 2015; Peters et al., 2019). The viability of the electrode-neural interface (e.g., spread of excitation, neural health) was unknown and so could not be accounted for in the analyses. The automatic labelling algorithm did not account for eCAPs with double peaks and hence excluding them from the analysis. The nature of data sharing in these analyses has made it possible to pool results from multiple data extraction experiments but for these retrospective data set background factors for individuals was unavailable and there was no control of recording settings at individual sites, however default approaches were used as recommended by the individual manufacturer. Despite all the limitations that would have increased the noise in the dataset, a weak relationship emerged to suggest that the lower the CF, the longer the n1p2 latency (r = 0.09, p = 0.024).

### Implications

The status of ENI can provide important information to better re-map Cls clinically (Lien et al., 2025). Knowing the relationship between CF and n1p2 latency, we could complement other ENI measures. For instance, the CF of the stimulated SGCs could be inferred by the measured n1p2 latency. Coupled with other ENI measures, clinicians can identify the location with poor neural health more easily and act accordingly. In addition, knowing SGCs differ along the cochlea, modern CI coding strategy could leverage this characteristic and provide and extra layer of time-coded information appropriate for the neural function across the array in each individual. For instance, a rate-varying coding strategy might be able to stimulate different SGCs despite overlapping stimulation pattern due to the spread of excitation and this could be personalised to stimulation rates that are appropriate for each individual.

### Observing such Relationship in a Typical Hearing System

An acoustically evoked compound action potential could ensure that the exact stimulated SGC population is known, provided the acoustic signal is delivered to a typically hearing ear. However, in contrast to recording with a CI, the recording electrode would be further away from the SGCs. This could be overcome by measures lowering the impedance, using longer average time, and or better signal processing algorithms. A study with trans tympanic needle electrode reported that the recorded response became narrower and the latency was shorter with the higher CF (Eggermont, 1976). This pattern was attributed to differences in basilar membrane delay, which are absent with electrical stimulus. The similar relationship is observed in eCAP data despite the lack of basilar membrane delay differences, suggesting that additional mechanisms may be involved. As a result, the authors plan to explore this relationship further with typical hearing participants using this acoustically evoked compound action potential approach and stimulus that produce a smaller basilar membrane delay difference.

### Plan for Future

Future studies should focus on a better estimate of CF (E.g., imaging tools to identify electrode location, other ENI measures to identify the spread of excitation, using acoustical stimulation with typical hearing participants), along with better control of other factors that might affect n1p2 latency (E.g., other ENI measures, unified eCAP recording settings). For Cl studies, recordings near threshold and at a low stimulation rate could limit the effect of spread of excitation and neural health on the n1p2 latency. The origin of the relationship might be attributed to the difference in firing probability over time and/or differences in the unit response across cochlea. Future work using deconvolution-based approaches could help disentangle these contributions (Strahl et al., 2016). With better study design, the authors believe that this relationship between CF and SGCs will be shown confidently.

## Conclusion

From independently shared datasets, using an approximation of insertion depth based on average values, a weak but significant correlation (r = 0.09, p = 0.024) was observed between the n1p2 latency, a proxy of action potential duration, and the CF of human SGCs. Human SGCs with lower CF may repolarise more slowly than higher CFs; an outcome which is in line with findings reported in the animal literature.

## Data Availability

All data produced in the present study are available upon reasonable request to the authors

## Notes

### Competing Interest Statement

The authors have declared no competing interest.

### Funding Statement

J.L. was supported by a joint scholarship provided by the Ministry of Education, Taiwan and Cambridge Commonwealth, European, and International Trust. C.G. was supported by MRC Impact Acceleration Award No. G116517. DV was funded by Medical Research Council MR/S002537/1, NIHR Cambridge Biomedical Research Centre NIHR203312, and NIHR Programme Grant for Applied Research NIHR201608.

### Author Declarations

This manuscript analysed shared de-identified data obtained upon request from several published studies. Each study obtained appropriate ethic approval at the time of experiment.

